# Profiling and Predicting Faculty Assessment Behavior in Surgical Education

**DOI:** 10.1101/2025.01.31.25321466

**Authors:** Ranish K. Patel, Phillip D Jenkins, Emily Lee, James Nitzkorwski, Ramanathan Seshadri, Rebecca Rhee, Mackenzie Cook, Julia Shelton, Julie Doberne, Jonathan Jesneck, Ruchi Thanawala

## Abstract

**Objective:** To study and profile the digital assessment behaviors of surgical faculty and residents, and to build a classifier to predict assessment completion, enhancing formative feedback initiatives.

**Background:** As competency-based paradigms are integrated into surgical training, developing digital education tools for measuring competency and providing rapid feedback is crucial. Simply making assessments available is inadequate and results in disappointingly low user participation. To optimize engagement and efficacy of these tools, user assessment behaviors need to be studied.

**Methods:** User data was aggregated from a HIPAA-compliant electronic health record (EHR)-integrated medical education platform. Faculty and resident behaviors were analyzed with respect to factors, such as time, day, device type, automated reminders, and EHR integration. A graphical convolutional neural network (GCN) model was trained to predict faculty participation in completing assessments.

**Results:** 10,729 assessments were completed by 254 attendings for 428 residents across 22 institutions, from 2022 to 2024. 86% of assessments were completed by faculty on weekdays, were significantly influenced by automated platform triggers and EHR integration, and distinct faculty behavior profiles contingent upon time to completion and comment length were established. Residents opened assessments at a median of 1.5 hours of faculty assessment completion, with 96% of assessments viewed by 24 hours. The GCN model successfully predicted faculty assessment completion with 93.5% accuracy and an area under the ROC curve of 0.97.

**Conclusions:** Faculty assessment behaviors represent an actionable bottleneck which can be leveraged to optimize and tailor the design of digital education tools, to enhance formative feedback.

## Introduction

Surgical residency and training is intended to prepare residents for safe, competent, and independent surgical practice. The competencies set forth by the Accreditation Council for Graduate Medical Education (ACGME) are high-level metrics assessing the global progress of a resident to meet this goal.^1^ Though, the day-to-day advancement of a resident toward practice-readiness occurs through frequent feedback and structured learning, feedback must be specific, consistent, and accessible to residents and faculty, and retrievable for future review.

Formative feedback is one of the most impactful processes in surgical training and adult learning.^2,3^ In an ideal educational environment, a resident would have access to formative feedback for every learning event, such as a consultation in the emergency department or a procedure in the operating rooms, and an opportunity to dialogue with the educator; however there are practical limitations to this practice, and it remains hard to do on a regular basis.^4-6^ While the challenges in providing regular feedback are multifactorial, variable teaching faculty practice habits contingent upon feedback timing is thought to be an important factor. For example, the time-frame of seventy-two hours following a clinical event has been established as the ideal time frame for the most effective feedback.^7^ This parameter has been embedded in educational tools and socialized as the optimal methodology.

The American Board of Surgery (ABS) Entrustable Professional Activity (EPA) program is driving the transition to a competency-based education (CBE) paradigm in surgical education. CBE requires high-volumes of data to understand baseline competency expectations across all trainees and programs.^8-11^ This high-volume data is also necessary to identify individual competencies and performance. Advancing this one-step further, precision medical education (PME) is predicated upon highly specific feedback and tailored education recommendations to personalize individual learning.^12,13^ Greater learner-specific granularity requires more data. CBE and PME rely on digital education tools (DETs), which can significantly increase formative feedback instances and store them electronically for retrieval. Additionally, tailoring DETs can optimize the various technology practice profiles of faculty conducting assessments.

We know little about surgical resident and teaching faculty DET user behaviors. Few studies have been dedicated to understanding user experience, behavior metrics and profiles. Most DETs have databases storing user behavior metrics, such as login times, platform or tool use time, content accessed, in addition to scores on performance assessments and recording feedback.^14^ This data is a rich source to study the usage behaviors of learners and educators and develop optimization strategies.

The field of human-computer interaction (HCI) is focused on optimizing the intersection of human psychology and computer system design. Understanding the workflow and experiences of users is essential in HCI. The ultimate goal of HCI optimization is the development of user-friendly, efficient, and practical technologies.^7,15^ HCI principles are useful to maximize the efficacy and impact of surgical DETs. As DET utilization expands within the realms of surgical education, identifying and understanding specific HCI trends and characteristics are of paramount importance, as they can inform and shape the technology used in order to improve the quality of feedback generated.

Prior research, now 10 years old, postulated that assessment quality decreases after 72 hours from time of event^7^. Taking into consideration the increased pervasiveness of mobile phones, web and app-based assessments, and the integration of digital solutions in modern surgical education, these data surrounding decay of assessment quality need to be reassessed.

In the present study, we sought to utilize a high-fidelity, electronic health record (EHR) integrated, HIPAA-compliant, web-based medical education platform to record user behaviors and to identify and define HCI trends in the facility-resident assessment interactions. We hypothesized that there are unique human behavior trends in faculty assessors and surgical trainee recipients of assessment results. Understanding these trends will help advance our knowledge of surgical DET user behavior, further develop DETs to include all user types and optimize their experiences. The ultimate goal is to improve the quality and quantity of assessments and feedback for our surgical residents.

This study evaluated faculty assessment behavior to understand why frequent assessments are difficult to reliably obtain, and we created a faculty assessor predictor model as a tool to help program directors understand which faculty will participate.

## Materials and Methods

For large-scale data collection, a HIPAA-compliant medical education platform was implemented at the study institutions to support surgical education information management (Firefly, www.fireflylab.org). The platform supports clinical and operative skill assessments, case assignments, automated case logging, and statistical inference of competency levels.^16,17^ It regularly synchronizes with the operative schedule from the various electronic health record systems at the individual institutions, providing case details, such as the date of operation, assigned staff and residents, assigned room, and procedures. For workflow flexibility, it supports multiple devices for synchronized data across computers and phones.

### Study and assessment design

Our study spanned from November 2022 to July 2024 and included attendings and residents from 22 institutions across the United States. At each institution, individual users were assigned accounts. Where EHR integration was established, schedules and case logs were auto-populated with their historical and future operations (Figure 1a). These data were derived from institutional operating room (OR) schedules, or from manually entered operations where OR schedule data was not available. This log was cumulative, maintained longitudinally, and it supported case logging assistance into ACGME and other logs. For each case, the platform considered the case details, such as procedures in operation, staff assignments, and resident training program type, and it used adaptive fuzzy matching algorithms to automatically match the case to the appropriate workplace-based assessment (WPBA). Although the platform provided several assessment types, for the broadest analysis across hundreds of surgical procedure types, this study focused on Component Assessment of Knowledge and Entrustability (CAKE) assessments (Figure 1b). These consist of procedure-specific operative questions paired with general skills questions. The procedure components were defined by a set of surgeon domain-experts within their surgical specialties. The assessment rating scale used an autonomy-based scoring system based on the Ottawa O-Score.^18^

**Figure 1:**
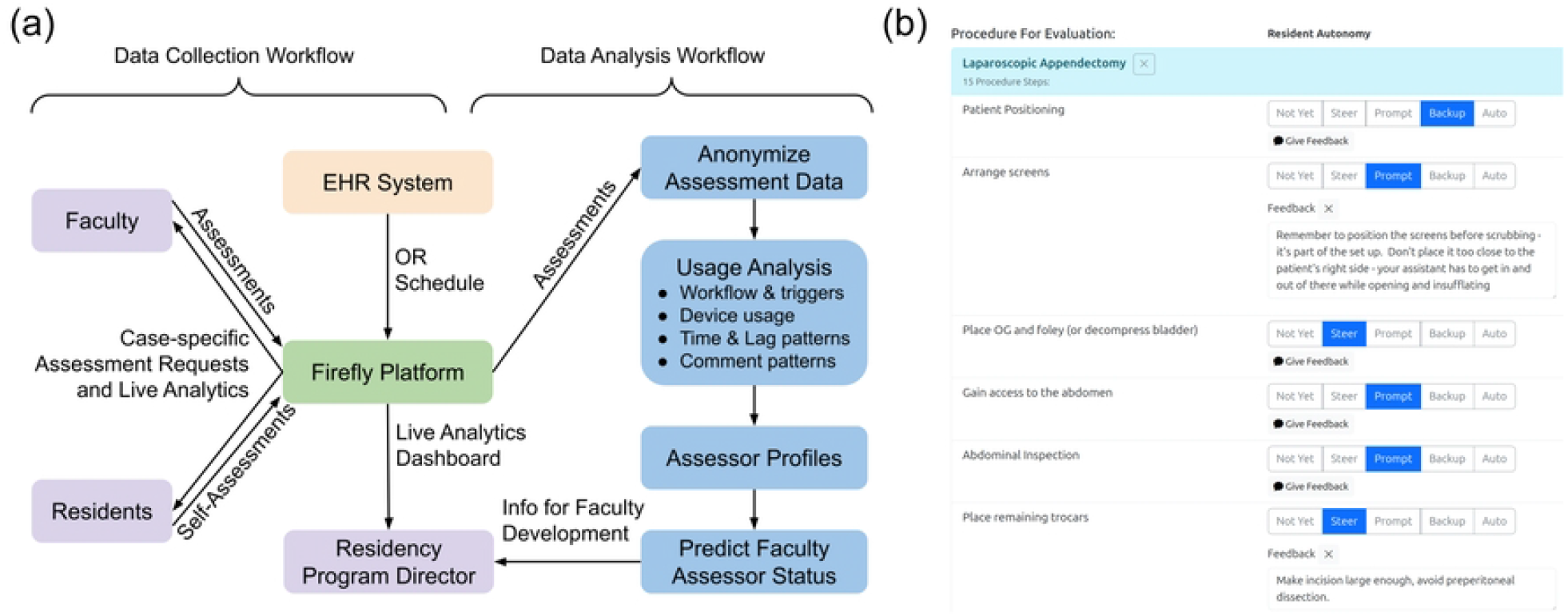

### Assessment collection

By identifying teaching cases from the OR schedule and case assignments, the platform automatically generated relevant WBPAs and prominently displayed them on an evaluations page to the faculty. To maximize the amount of feedback for residents and encourage faculty participation, the platform automatically requested WPBAs from the faculty at 1700 local time on weekdays. This time was chosen to present the assessment task to the faculty member during a quieter part of the work day, when it would be more convenient to complete an assessment.

However, this was a default notification setting, and users could set their own notification preferences as desired. Whenever a WPBA was completed, the platform immediately notified the resident and sent a link for the resident to click to open the assessment and review the feedback. All completed assessments were available for review by the completing teaching faculty and the recipient resident in perpetuity. The assessment results were incorporated into an analytics dashboard to allow for dynamic review of performance.

### Exploratory analysis of assessment behavior

In order to explore factors that contribute to faculty assessment habits, the platform captured various user-specific metrics, including faculty procedure types, case start time, the frequency of operating with residents, user device type (computer vs. phone), WPBA completion timestamps (date and time), and the resident WPBA review timestamps. For this study, we aggregated and analyzed the raw data from the Firefly platform database along with the user activity logs. For exploratory analysis, we calculated the cumulative WPBA data volume accrued over time, and tallied the counts of WPBAs completed over the days of the week and the time of day. Faculty assessment behavior preference was calculated with Chi-squared tests by considering WPBA counts according to various factors, including weekdays vs. weekends, computer vs. phone as the device, in institutions with vs. without EHR data integration, and by workflows of initiating the WPBA manually vs. from automated triggers based on the OR schedule.

### Lag time analysis

In order to assess the effect of time lag after cases, the WPBAs remained available for completion for two calendar weeks (14 days) from the date of the operation. If the faculty did not fill out the WPBA in this timeframe, the platform then automatically expired the WPBA, removed it from the faculty’s queue, and marked it as “not completed” in the platform’s database. We also considered the effect of lag time on the presence of WPBA comments and comment length.

Similar to faculty assessment lag time, we also investigated resident WPBA open lag time. The open lag time was defined as the time between the WPBA being delivered to the resident (immediately upon faculty completion) and the time when the resident opened the WPBA. This open lag provided inference to resident attitudes about the value of the feedback.

### Predicting faculty assessor status

In addition to exploratory analysis, we were interested in testing whether the assessment behavior data enabled us to predict the assessor status of each faculty member. Predicting whether faculty participate in completing WPBAs is valuable information for a residency program director, for managing educational initiatives, optimizing the quality of feedback going to residents as learners, and for faculty development initiatives. Therefore we utilized the platform’s predictive model that can learn from interpersonal interactions in social-network type data, a graphical convolutional neural network (GCN)^19-21^. The model inputs consisted of case patterns between individual faculty members and residents operating together, faculty institution, department, and surgical procedure types. The GCN output was the probability of a faculty member being an assessor (participating in completing any assessments). The GCN model was trained on a high-performance cluster of servers with dedicated graphics processing unit (GPU) hardware for machine learning computations on the Amazon Web Services (AWS) system. To prevent overfitting to the data, the GCN model was trained and tested with 10-fold cross validation. To quantify the model’s performance, we used the metrics of accuracy, area under the receiver operating characteristic (ROC) curve, precision-recall curves, and the F-score.

## Results

Across 22 institutions, 254 faculty members completed 10,729 CAKE assessments for 428 residents over the study period of November 2022 to July 2024. Throughout the study period, there was a continuous rise in the number of CAKE assessments completed, with a year-over-year growth rate of 169% (Figure 2a). Faculty assessments were more commonly completed on weekdays (86% on weekdays vs. 14% on weekends, Chi-squared test p-value <0.00001) (Figure 2b). Weekend assessment completion demonstrates that faculty did develop persistent assessment completion habits without reminders. Faculty completed a median of 18 assessments.

**Figure 2:**
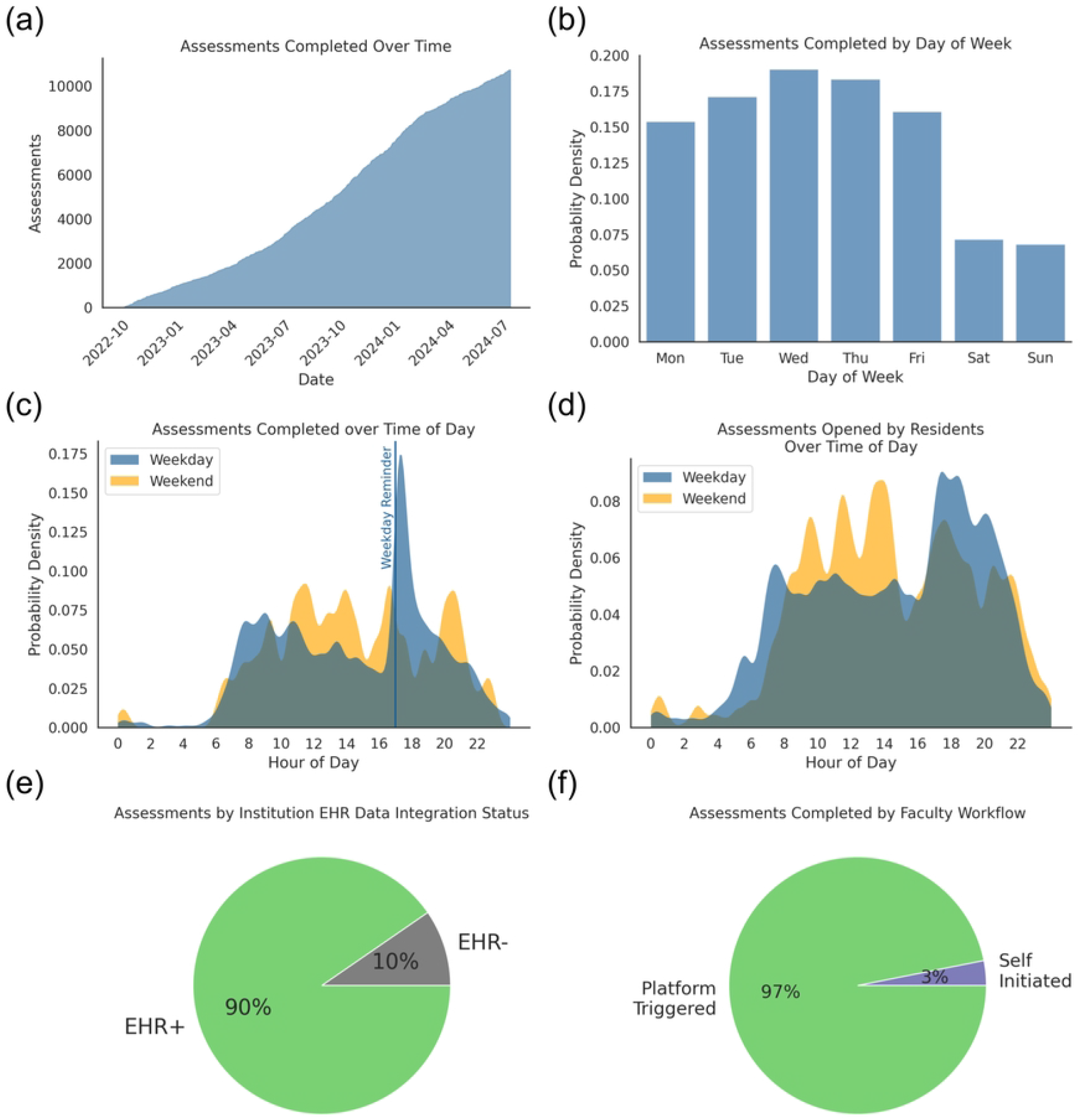

For time of day, faculty showed a significant spike in completing assessments after the 5 pm daily reminder, and otherwise generally preferred the work mornings (7:30 - 10:00 am) (Figure 2c). Faculty continued this habit over the weekend, but shifted later in the day by approximately 3 hours. Residents generally opened their assessments as soon as practical upon receiving them (Figures 2d and 4b). Workflow analysis showed that the most significant factor supporting assessment completion was the institution’s EHR integration status with OR schedule data.

EHR integration resulted in 9.4-fold increase in assessment completion (90.4% in EHR+ vs. 9.6% in EHR-institutions; p-value < 0.00001, one-way Pearson Chi-squared test) (Figure 2e). Platform triggered alerts, derived from OR schedule data, for assessment completion dramatically drove faculty behavior, with 97% of assessments completed via a platform trigger versus only 3% being self-initiated (p-value < 0.00001, one-way Pearson Chi-squared test) (Figure 2f).

Faculty presented a wide spectrum of assessing behaviors (Figure 3). Behavioral patterns fit power law distributions, especially with early adopters diving into frequent platform usage, and most other faculty following along thereafter. Faculty as a whole completed assessments for a median of 7 different procedure types, assessed a median of 6 unique residents, and residents were assessed by a median of 5 unique attendings (Figure 3a, 3b, 3c). Faculty assessing behavior was then characterized by several factors. Faculty were sorted along a spectrum of specialist to generalist assessors, by comparing the number of residents assessed and over the number of procedure types (Figure 3d). Considering the lag time between cases and their associated assessments, faculty were grouped into early, middle, late, and variable lag assessors (Figure 3e), and then their assessment comment behaviors grouped the faculty by comment length and frequency (Figure 3f).

**Figure 3:**
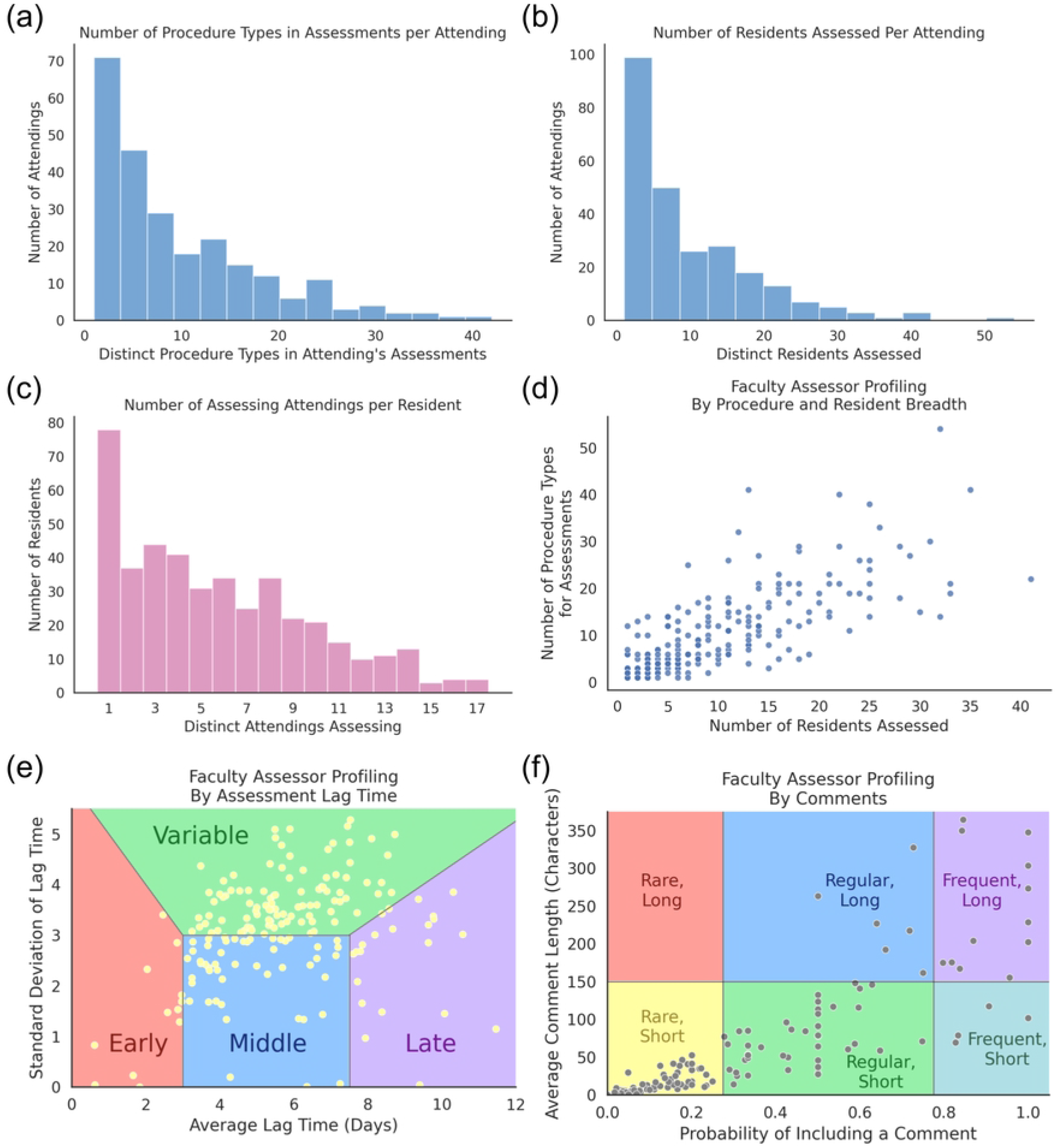

The assessing lag time was measured between a case and its associated assessment, and the peak assessment activity occurred within 24 hours of the case. Faculty completed assessments with a median lag time of 3.42 days after the case start time (Figure 4a). There was a gradual decrease in the proportion of assessments completed sequentially out to 1 week after the date of the operation. Whereas faculty often waited to complete assessments, residents opened their assessments rapidly after receiving them. Residents opened most assessments to review results within 5 hours of completion by teaching faculty (median of 1 hour 30 minute lag time), including assessments received late at night during sleeping hours. Almost all assessments (96%) were reviewed by 24 hours (Figure 4b).

**Figure 4:**
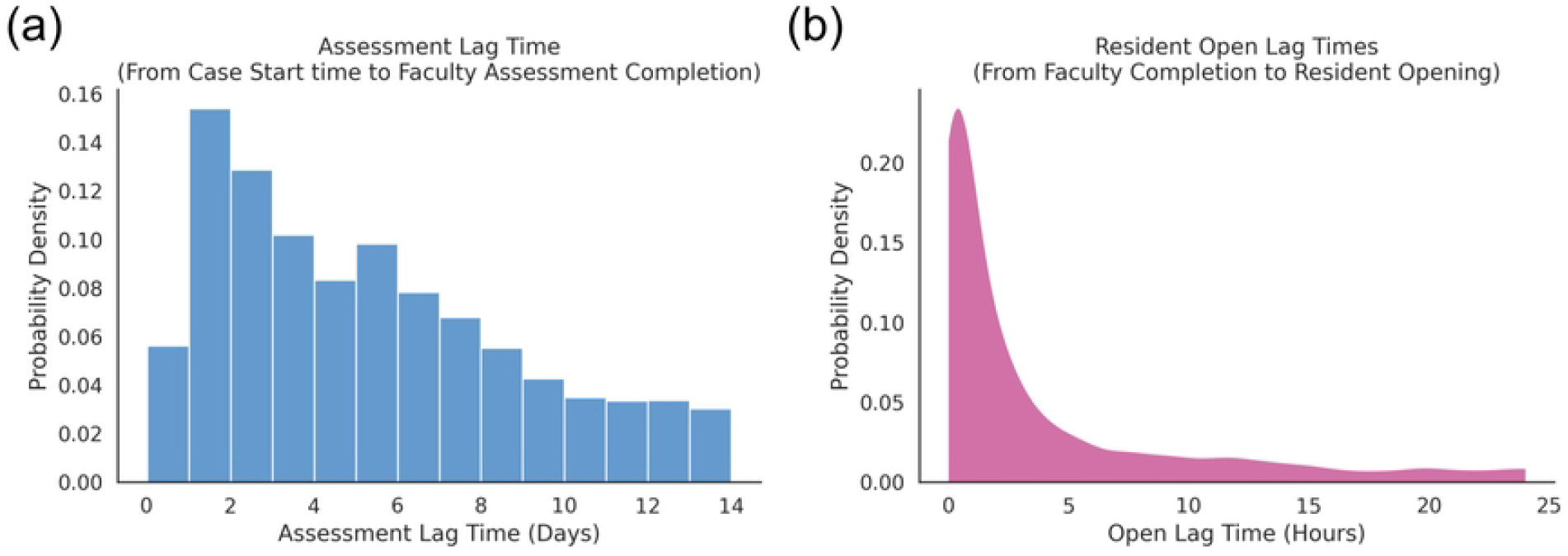

The Firefly platform was available on multiple device types, and both faculty and residents displayed preferences for device usage. Faculty as a whole had a strong preference for using a computer, with 69% of assessments completed on a computer vs. 31% on a phone (p-value < 0.00001, Chi square test) (Figure 5a). Although Faculty preferred using a computer overall, there were periods of increased phone usages in the busy workday morning and also in the evening after the 5 pm reminders (Figure 5b). For reviewing the assessments, residents as a whole showed balanced usage across computers and phones. However, individual faculty members and individual residents showed very strong device preference, with few individuals showing flexibility in usage across multiple device types (Figures 5c and 5d).

**Figure 5:**
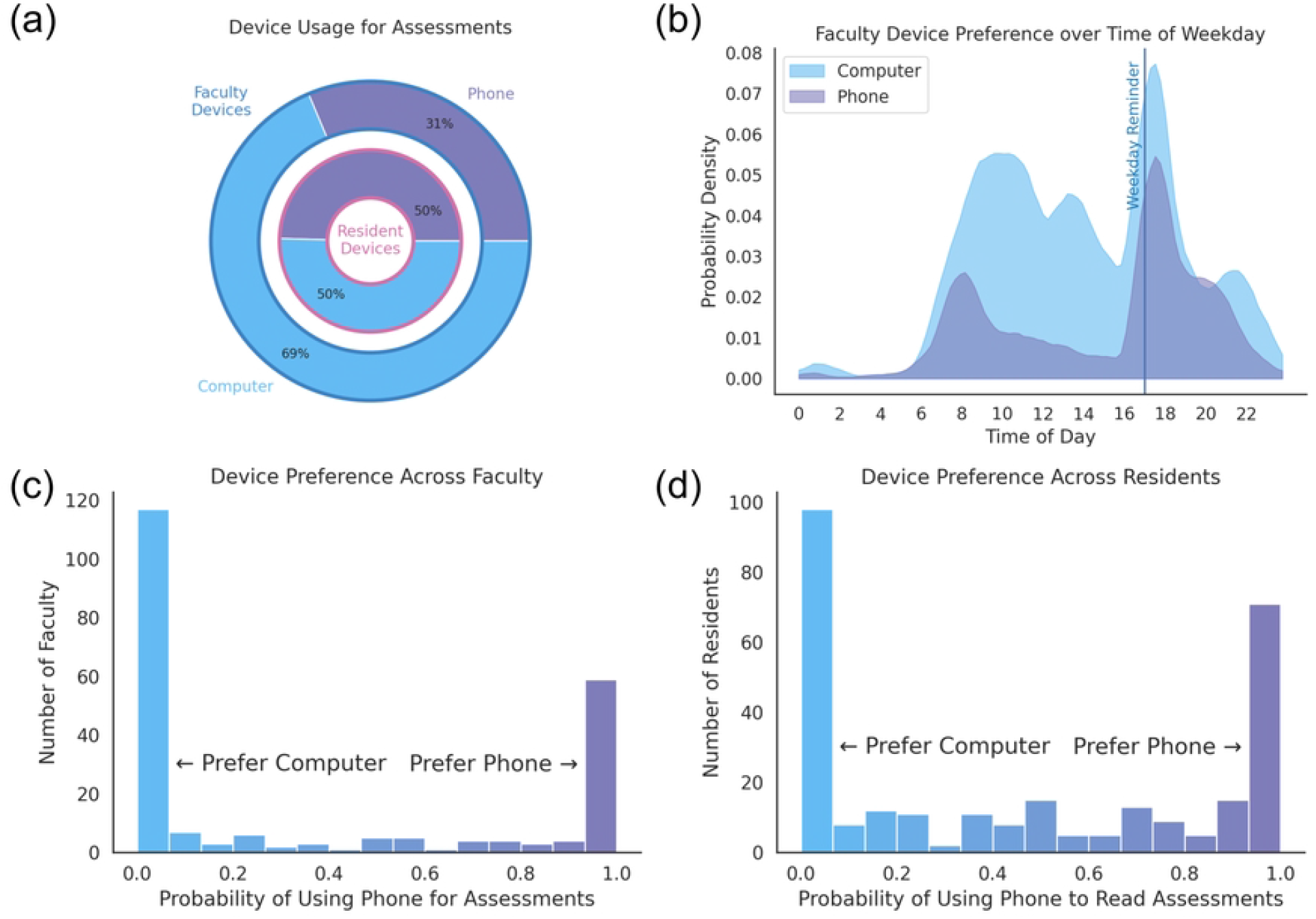

Device preference showed follow-on effects on assessment data entry, most noticeably for writing assessment comments. Despite the option to dictate comments on a phone, faculty preferred to write comments on a computer, including comments on 19% of assessments on a computer but only 14% on a phone (p-value < 0.00001, Chi square test) (Figure 6a). The comments by computer also tended to be more detailed. Faculty wrote longer comments on a computer, with a median comment length of 153.0 characters on a computer and 129.5 on a phone (p-value < 0.00001, T-test with unequal variances) (Figure 6b). Comments also were the largest factor determining the assessment completion time. Faculty completed the assessments very quickly, with a median time of 27.7 seconds overall, split into 97.7 seconds with comments vs. 22.1 seconds without comments (p-value < 0.00001, T-test with independent samples) (Figure 6c).

**Figure 6:**
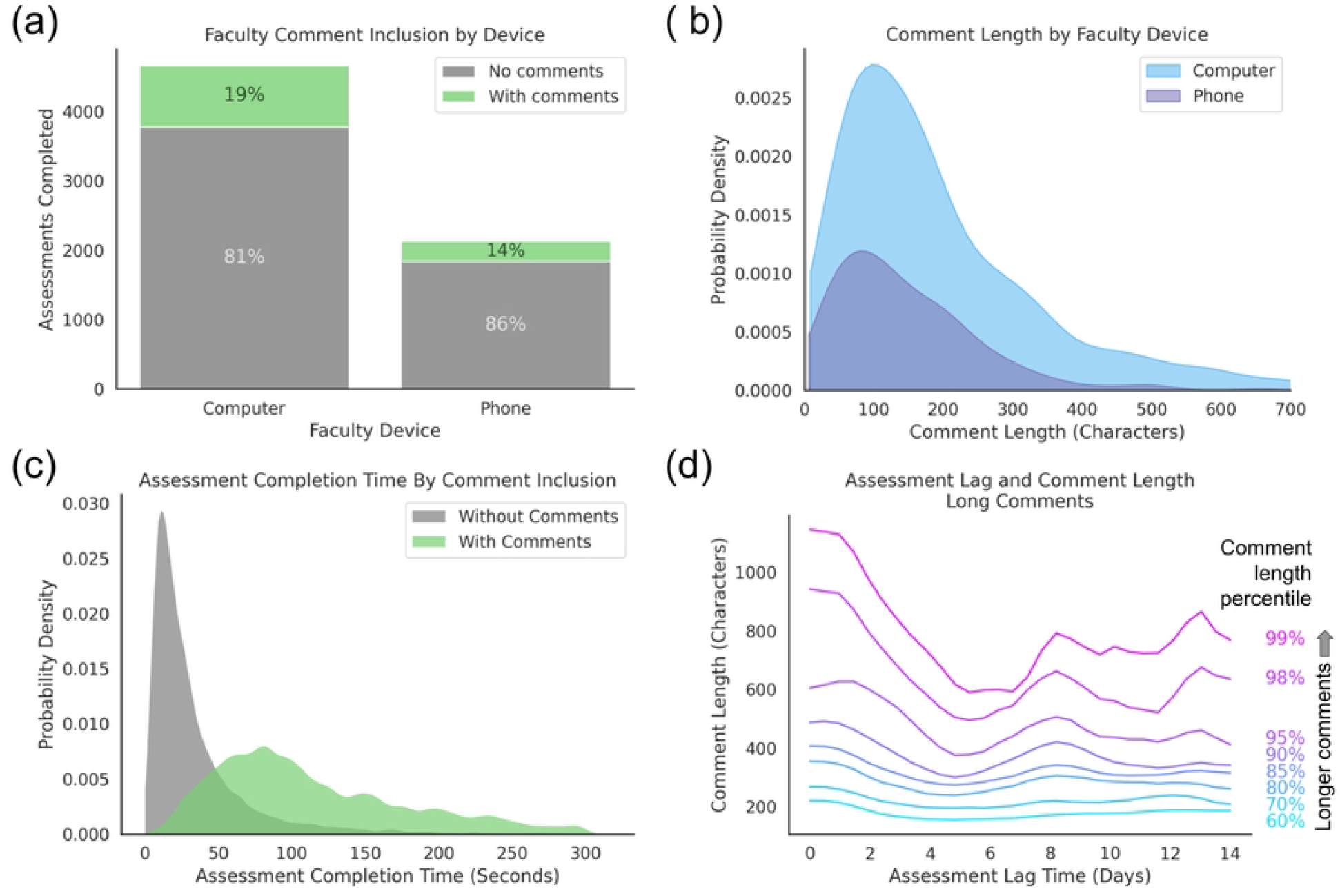

Comment length also showed an interesting bimodal distribution pattern with lag time (p-value = 0.021, Hartigan’s dip test) (Figure 6d). Maximum comment length occurred within the first 24 hours after a case, and most detailed comments occurred in early assessments within the first couple days. Then after a dip towards the end of the first week, a second peak of longer comments was seen at a 1 week lag. This trend was consistent for both short and long comments, but more pronounced for long comments.

In order to predict the probability of each faculty member being an assessor (completing at least one assessment), we compiled operative data from surgical OR schedules and then validated the case assignments with resident case logs. The operations showed co-operating patterns between faculty and residents (Figure 7a). To augment the case assignments, we added relevant user data, such as the faculty’s institution, department, most common procedure types, and details of their teaching cases with residents. For predictions, we used the platform’s graphical convolutional neural network model. Trained with 10-fold cross-validation, the network model predicted the faculty’s assessor status with near-perfect performance (accuracy of 0.935, F1 score of 0.828, an area under the receiver operating characteristic (ROC) curve of 0.97, and an average precision of 0.90) (Figure 7a and 7b).

**Figure 7:**
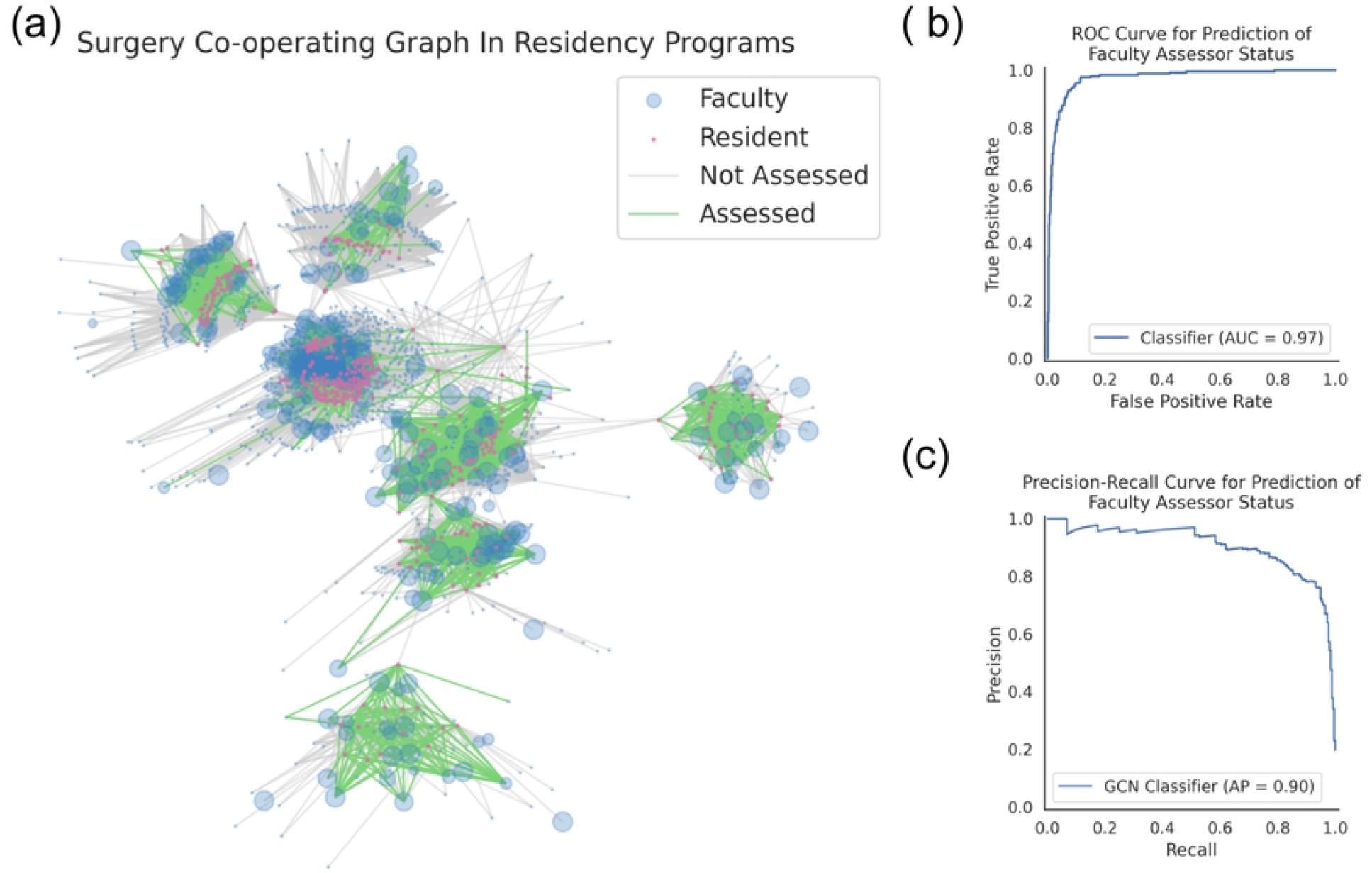

## Discussion

CBE models require frequent assessments of residents for success and learner-specificity.^22-24^ It is paramount to develop and implement high-quality DETs that facilitate these assessments.

These tools need to be flexible and inclusive of all user behaviors to maximize user participation. Human-centered design is dependent on co-evolution of a technology with user needs.^25^ “Build it and they will come” is an abolished concept in technology development.^26^ Understanding what each user group needs is essential to the successful iterative development of any new tool. Workflow mapping is a key principle in informatics and a critical primary step to ensure that no valuable user is left behind.^27^ It is an often overlooked step as we focus on developing the core functionality of a technology that fills a known gap; assessments in this case. But without workflow mapping and subsequent study of user patterns, we are unknowingly excluding critical data on our resident learners. We describe in detail the various use and profile patterns of teaching faculty and residents using our multi-institutional, EHR-integrated DET.

Automated assessment matching and user reminders driven by EHR-derived data eliminated the need to remember to complete assessments. This functionality captures assessments that would not have been completed otherwise in a manually initiated process.^9^ With a maximally inclusive and complete dataset on assessments and user patterns, we map clear trends and profile characteristics in residents and teaching faculty. As shown in our analysis, there is a 9.4 fold increase in assessment completion with EHR integration. The impact of reminders on faculty was even more drastic, with 97% completing their assessment after the reminder was sent.

Residents and teaching faculty have distinctly different behaviors regarding assessments. Residents are consistently rapid consumers of data about their performance. On the other hand, teaching faculty are highly variable on when they complete assessments, the inclusion and length of their comments. Assessment availability for 14 days after an operation or procedure allowed us to demonstrate that assessments are completed between 7 to 14 days after the clinical event for a late group of teaching faculty. This group has been historically excluded with the more restrictive 72 hour assessment timeframe guidance.^7^ The 72-hour limit was likely needed due to the manual completion of the Operative Performance Rating System assessments (OPRS). The DET in this study maintains contextual details of the operation, likely improving recall for the teaching faculty on details of resident performance.

Detailed assessments occur at two timepoints, within 24 hours and about 1 week later. This bimodal distribution likely reflects the dual-process theory of memory and learning, system 1 and system 2. System 1 processing is defined by fast, unconscious recall of information. In learning theory, this would be regurgitation of memorized information. System 2 processing is used when critical analysis, applications of less-used concepts, or novel solutions are needed. System 2 processing takes more time compared to system 1 processing. It is plausible that this second peak at 1 week post-clinical event reflects system 2 type processing of resident performance. DET flexibility to include teaching faculty assessor who are system 2 processors is important as these may hold some of the most formative comments. For future work, we are examining comments through language analysis using computational tools such as computational linguistics and natural language processing of assessment comments.

Preference for computer use over mobile devices holds true this study of our CAKE assessment dataset similar to our previous work in EPA user behaviors.^9^ The cognitive load for assessment completion across both types of assessments is at a threshold where mobile devices, even with in-built dictation tools, are felt to be inadequate by most teaching faculty. Flexible device support is a necessary quality of a modern DET tool.

Even with the many flexibilities built into the DETs used in this study, our network model demonstrates that there is “cherry-picking” of operations where assessments are completed. Also, not all teaching faculty are assessors. There are identifiable patterns allowing us to classify individuals as assessors and non-assessors with a high-degree of confidence and model performance. How do we convert the non-assessors to assessors? Why are non-assessors not doing assessments? There remain hidden factors to be examined to answer these questions. But, the answers can be found through the study of user behavior in well-designed DETs.

The granularity of information to specifically understand how our residents are learning and how our teaching faculty are driving learning requires capture and inclusion of every assessment possible. We cannot assume that developing assessments without careful examination of how we are completing them, the implementation processes, and the ongoing user perceptions, utility and behaviors will be enough in our evolution to CBE.

## Conclusions

Human-centered design is essential in DETs for the iterative development and evolution of these technologies to capture all possible data on resident learning. EHR integration and automated reminders are key drivers in assessment completion by faculty. Teaching faculty have diverse user profiles that should be accommodated for in DETs. High-value feedback can occur 1 week after a clinical event likely due to the different types of memory processing by teaching faculty. Network modeling can reliably predict faculty into assessors versus non-assessors. Computational analysis of user log data and behaviors has provided insights that should be incorporated to future iterations of DETs. Adoption of EHR integrated DETs has shown dramatic increases in assessment completion, and allowed for unparalleled analysis of specific faculty behaviors that are the drivers of learner assessment, and the future of CBE in surgical education.

## Data Availability

The data is premised on learner and hospital-based data which has been completely de-identified. Data governance policies of the research group require knowledge the individuals and intention of how the data would be used before sharing the de-identified datasets used in this study

